# Continued Age Shift of Confirmed Positive COVID-19 Incidence Over Time to Children and Young Adults: Washington State March - August 2020

**DOI:** 10.1101/2020.11.12.20229468

**Authors:** Judith Malmgren, Boya Guo, Henry G. Kaplan

## Abstract

**Background:** As the coronavirus (COVID-19) epidemic passed the initial infection peak in Washington State, phased re-opening lifted stay-at-home orders and restrictions leading to increased non-essential work, social activities and gathering, especially among younger persons.

**Methods:** A longitudinal cohort analysis of Washington State Department of Health COVID-19 confirmed case age distribution 1) March-April 2020 (N=13,934) and 2) March-August 2020 (N=76,032) for proportional change over time using chi square tests for significance.

**Results:** From March 1^st^ to April 19, 2020 COVID-19 case positive age distribution shifted with a 10% decline in cases age 60 years and older and a 20% increase in age 0-19/20-39 years (chi-square = 223.10, p <.001). Number of cases over the eight-week analysis period were 0-19 years n = 515, 20-39 years n = 4078, 40-59 years n =4788, 60-79 years n = 3221, 80+ years n = 1332. After the peak (March 22, 2020), as incidence declined in older age groups, the combined percentage of cases age 0-19 and 20-39 increased from 20% to 40% of total cases. During this time testing expanded with more testing among older age groups while case positivity shifted young. Percent positive cases by age through August 2020 increased to a consistent average of 60% less than age 40 [age 0-19 increased to 19% (N = 10257), age 20-39 increased to 42% (N = 30215)].

**Conclusions:** An increased share of COVID-19 incidence among children (age 0-19) and young adults (age 20-39) indicates their elevated role in propagating the epidemic by creating a reservoir of disease with risk of spillovers to more vulnerable older persons and those with comorbid conditions. Media savvy and age appropriate COVID-19 messaging may increase mitigation compliance among these less vulnerable, more mobile and lower priority vaccination age groups. As vaccines become available, mitigation will continue to be a priority to reduce overall population incidence.

## Introduction

The 2019 novel coronavirus (COVID-19) has quickly spread over the globe and positive cases continue to climb. The first reported case of 2019-nCoV infection in the United States was in Washington State on January 19, 2020 and on March 23^rd^, a statewide stay-at-home order was announced to remain in effect until May 4^th^ which was extended to May 30, 2020 and then lifted in a move to phased reopening.^1,2^ Change of stay-at-home restrictions, social gathering guidelines and reopening of businesses lead to an increase of non-essential work and social activities, especially among younger persons. Hospitalization and death rates associated with older age and comorbid conditions declined within a short time after stay at home orders were issued. These rates continued to decline over time while the number of confirmed positive cases plateaued, indicating shift to younger age of incident cases if present could explain the changed presentation and severity of cases.^3^ Using Washington State Department of Health (DOH) data, we analyzed incidence of COVID-19 cases by age over time for significant change in age distribution as the pandemic progressed.

## Methods

Testing of symptomatic patients for COVID-19 in Washington State steadily increased since the outbreak began in January 2020 with drive-through testing beginning in some communities in late March increasing testing availability. Testing sites continued to increase into the summer months. More than 25 labs in the state are now able to process COVID-19 testing. Total testing capacity statewide continues to change, which means each lab is the best source for current numbers on their own testing capacity. The results from all COVID-19 tests flow into the Washington Disease Reporting System (WDRS), an electronic disease surveillance system that allows public health staff in Washington state to receive, enter, manage, process, track and analyze disease-related data. Aggregate testing data is updated and published daily on the WA State DOH website with an update by age and county weekly.

For the initial data analysis and trend reporting, weekly updated COVID-19 positive confirmed case data from January 16^th^ to April 19^th^, Washington State DOH were used which are available to the public on the DOH website.^4^ The updated data for trend over time in age distribution for weeks 4/26-8/23/2020 was obtained from the same source. Lab-confirmed COVID-19 cases from hospital, intensive care units, emergency departments and outpatient testing are reported to DOH by acute care hospitals in Washington daily. The data from DOH includes the number of cases, deaths and hospitalizations by week, county and age groups. Additional publicly available data on number of tests performed was also used in our study to address possible bias of changed testing by age over time at the beginning of the pandemic. Our analysis for age trend over time used data through the week of 8/23/2020 to stay in the time frame when schools were still closed, updated September 20, 2020 to allow for a time lag in reporting.

Laboratory-confirmed county-assigned cases by state assigned age groups 0-19, 20-39, 40-59, 60-79 and 80+ years in Washington State were enumerated and plotted over time. Statistical analyses were performed using Microsoft Excel for percentage calculations, an online chi square calculator for two-sided significance testing with a .05 level of significance and an online chi-squared test for trend.^5,6,7^ Weeks for the first analysis were restricted to March 1 to April 19 2020 when a sufficient number of cases (20 or more) had accrued to be statistically relevant and to accommodate the two-week lag time in case reporting and confirmation to WDRS. Age 0-19 was included even though there were only 4 cases in the first analysis week but case counts were greater than 20 in following weeks. Chi-square tests were run comparing the first two weeks of increased cases (3/1/2020, 3/8/2020), the peak week (3/22/2020) and weeks (4/12/2020, 4/19/2020). Initial comparison weeks were restricted to 3 groups and time from 3/1/2020 to 4/19/2020 to avoid the diminishment of statistical validity by multiple comparisons. Chi square statistics were calculated for testing by age to the same initial time period to assess the degree to which age group testing was equivalent to case positive distribution or over sampling was done by age group. The secondary analysis was a chi square test for slope (linear trend) of percentage confirmed positive cases age less than 40 using the last weeks of April through August for significant additional age shift from older to younger cases over time.

## Results

Total number of positive cases in Washington State reported from January 16^th^ to April 19^th^ excluding positive cases with unknown age (n=10) were 14,220 with 13,934 confirmed cases from March 1 to April 19 [0-19 = 515 (4%), 20-39 = 4078 (29%); 40-59 = 4788 (34%); 60-79 = 3221 (23%); 80+ = 1332, (10%)]. The four counties with the highest number of cases by rank order were King (n=5955), Snohomish (n=2300), Pierce (n=1300), and Yakima (n=1119). As the epidemic progressed and the curve flattened, fewer older and more young persons tested positive for COVID-19 with the percentage of total cases among age 0-19 and 20-39 doubling from 20% to 40%. Figure 1. There was an increase in cases age 0-19 years over time from four cases week 3/1/2020 to 83 cases week 4/19/2020 and 131 cases week 5/3/2020. Incidence among age 60 and older declined 55% off peak but 20-39-year-old cases only declined 36% off the peak week (3/22/2020). Table 1. The chi-square test statistic for the comparison of confirmed cases by age across the three discrete time periods of weeks 3/1-3/8, 3/22, and 4/12-4/19, was 223.10, p<.001. Table 1.

**Table 1.** WA State DOH COVID-19 confirmed cases by age: 3/1/2020-4/19/2020 (n=13934)

|  |  |  |  |  |  |  |  |  |
| --- | --- | --- | --- | --- | --- | --- | --- | --- |
|  | 0-19<br>N (%) | 20-39<br>N (%) | 40-59<br>N (%) | 60-79<br>N (%) | 80+<br>N (%) | Row<br>Total | Chi<br>Square | p-value |
| Week |  |  |  |  |  |  |  |  |
| 3/1* | 4<br>(.7%) | 111<br>(19%) | 184<br>(31%) | 209<br>(36%) | 78<br>(13%) | 586<br>(4%) | 223.10 | <.001 |
| 3/8* | 37<br>(2%) | 382<br>(25%) | 518<br>(33%) | 449<br>(29%) | 165<br>(11%) | 1551<br>(11%) |  |  |
| 3/15 | 38<br>(2%) | 680<br>(30%) | 836<br>(37%) | 559<br>(24%) | 178<br>(8%) | 2286<br>(16%) |  |  |
| 3/22* | 65<br>(3%) | 708<br>(28%) | 917<br>(37%) | 548<br>(22%) | 265<br>(10%) | 2509<br>(18%) |  |  |
| 3/29 | 101<br>(4%) | 669<br>(30%) | 743<br>(33%) | 554<br>(23%) | 226<br>(10%) | 2258<br>(16%) |  |  |
| 4/5 | 86<br>(5%) | 524<br>(29%) | 645<br>(36%) | 379<br>(21%) | 160<br>(9%) | 1794<br>(13%) |  |  |
| 4/12* | 101<br>(6%) | 550<br>(34%) | 516<br>(32%) | 315<br>(19%) | 139<br>(9%) | 1621<br>(12%) |  |  |
| 4/19* | 83<br>(6%) | 454<br>(34%) | 429<br>(32%) | 237<br>(18%) | 126<br>(9%) | 1329<br>(10%) |  |  |
| Column<br>total | 515<br>(4%) | 4078<br>(29%) | 4788<br>(34%) | 3221<br>(23%) | 1332<br>(10%) | 13934<br>(100%) |  |  |
| 4/26** | 131<br>(8%) | 542<br>(35%) | 531<br>(34%) | 219<br>(14%) | 131<br>(7%) | 1554 |  |  |
| 5/3** | 131<br>(11%) | 466<br>(39%) | 364<br>(30%) | 164<br>(14%) | 85<br>(7%) | 1210 |  |  |
| Column<br>Total** | 777<br>(4%) | 5086<br>(30%) | 5683<br>(34%) | 3604<br>(22%) | 1548<br>(9%) | 16698 |  |  |

\*\* Not included in the chi-square test analysis

**Figure 1.**
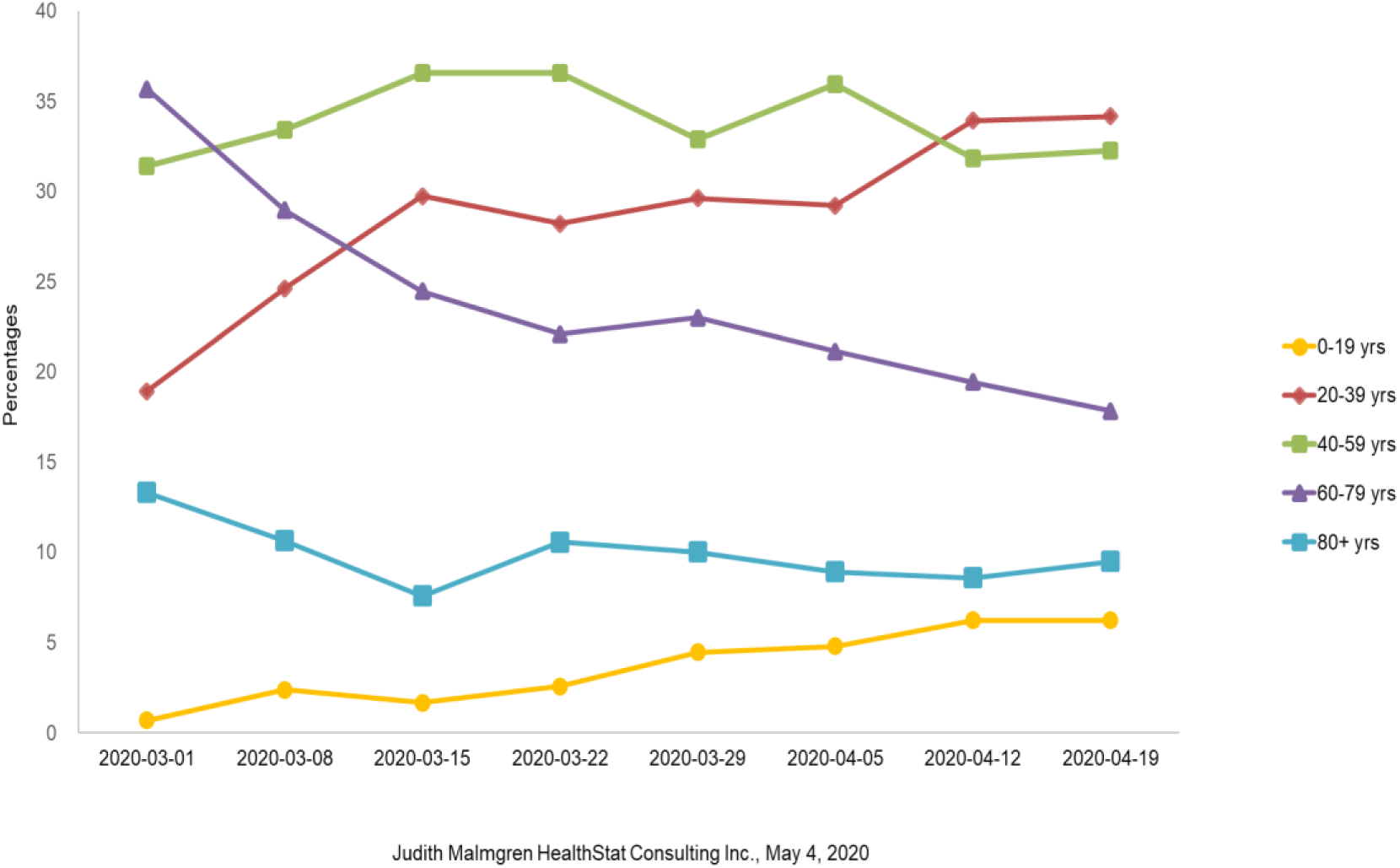
WA State DOH COVID-19 Confirmed Cases by Age: 3/1/2020-4/19/2020

A total of 186,655 Covid-19 tests were run during the March 1 to April 19 2020 time period with 7.5% positive over the eight-week time period. Percent positive testing was variable over time and ranged from 12% (3/1/2020) to 5% (4/19/2020). Age distribution of persons tested changed by a small margin over time with an increase in testing among persons age 60 and older (+6%) and a decrease in testing among 0-19 and 20-39-year olds (−3%) from the peak (3/22/2020) to the week of 4/19/2020, a statistically significant but small change [chi square = 263.87, p<.001]. Table 2, figure 2.

**Table 2.** WA State DOH testing by age group: 3/1/2020-4/19/2020 (n=184645)

| Table 2. WA State DOH testing by age group: 3/1/2020-4/19/2020 (n=184645) |  |  |  |  |  |  |  |  |
| --- | --- | --- | --- | --- | --- | --- | --- | --- |
| Week | 0-19<br>N (%) | 20-39<br>N (%) | 40-59<br>N (%) | 60-79<br>N (%) | 80+<br>N (%) | Total | Chi-square | P<br>value |
| 3/1 | 313<br>(16%) | 460<br>(24%) | 507<br>(26%) | 478<br>(25%) | 187<br>(10%) | 1945<br>(1%) | 263.87 | p<.001 |
| 3/8 | 2006<br>(13%) | 4911<br>(31%) | 4992<br>(31%) | 3162<br>(20%) | 863<br>(5%) | 15934<br>(9%) |  |  |
| 3/15 | 2231<br>(8%) | 9544<br>(35%) | 9041<br>(34%) | 5034<br>(19%) | 1098<br>(4%) | 26948<br>(15%) |  |  |
| 3/22* | 1832<br>(6%) | 10257<br>(36%) | 10000<br>(35%) | 5587<br>(19%) | 1177<br>(4%) | 28853<br>(16%) |  |  |
| 3/29 | 1825<br>(7%) | 9851<br>(35%) | 9611<br>(34%) | 5445<br>(19%) | 1331<br>(5%) | 28063<br>(15%) |  |  |
| 4/5 | 1681<br>(6%) | 9041<br>(34%) | 9235<br>(34%) | 5658<br>(21%) | 1273<br>(5%) | 26888<br>(15%) |  |  |
| 4/12 | 1612<br>(7%) | 7945<br>(32%) | 7807<br>(32%) | 5550<br>(22%) | 1753<br>(7%) | 24667<br>(13%) |  |  |
| 4/19* | 2094<br>(7%) | 9711<br>(31%) | 10572<br>(34%) | 7238<br>(23%) | 1732<br>(6%) | 31347<br>(17%) |  |  |
| Total | 13594<br>(7%) | 61720<br>(33%) | 61765<br>(34%) | 38152<br>(21%) | 9414<br>(5%) | 184645 |  |  |

**Figure 2.**
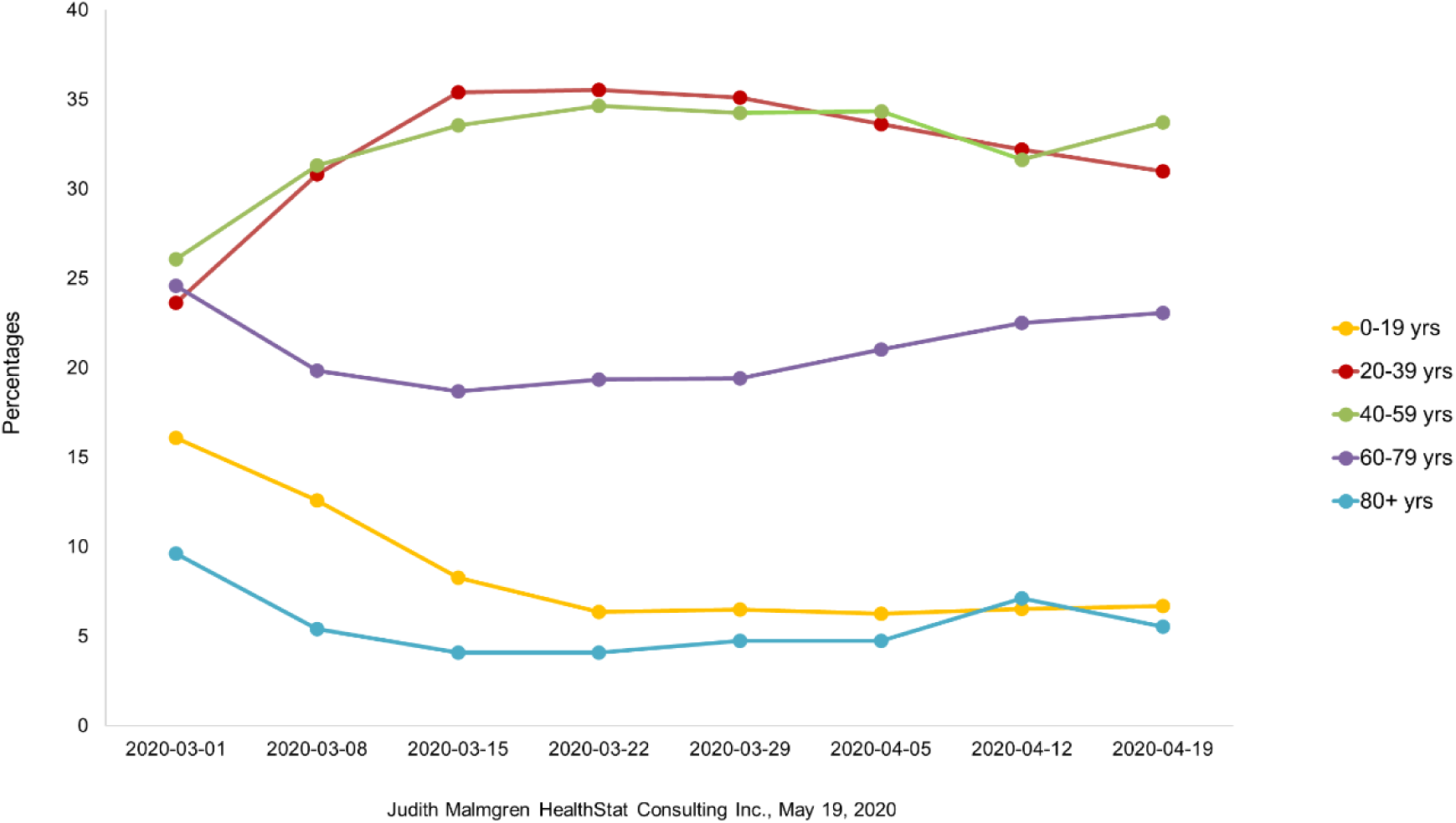
WA State DOH COVID-19 testing by age group: 3/1/2020-4/19/2020

From the first COVID-19 peak week (3/22/2020) to week 4/19/2020, the COVID-19 hospitalization rate declined 49% for age 40 and older cases with a 54% decline in COVID-19 incidence. During the same time period, the hospitalization rate among age 20-39-year-olds declined 35% with a 36% decline in COVID-19 incidence.^3^ Over the longer time period of April to August 2020 the shift to younger age of COVID-19 confirmed positive cases continued to a sustained average of 60% of cases age less than 40 years June-August 2020 (N=76,032). Figure 3. The chi-square test for trend from April to August 2020 using the last week of each month comparing age of confirmed positive cases less than 40 to age 40 and older was significant indicating the slope was not zero over time [chi-square = 82.97, p<.001]. However, the chi-square test for trend for May to August 2020 was not significant indicating the proportion of cases less than age 40 reached a constant plateau in those months [chi-square = 1.41, p=.24].

**Figure 3.**
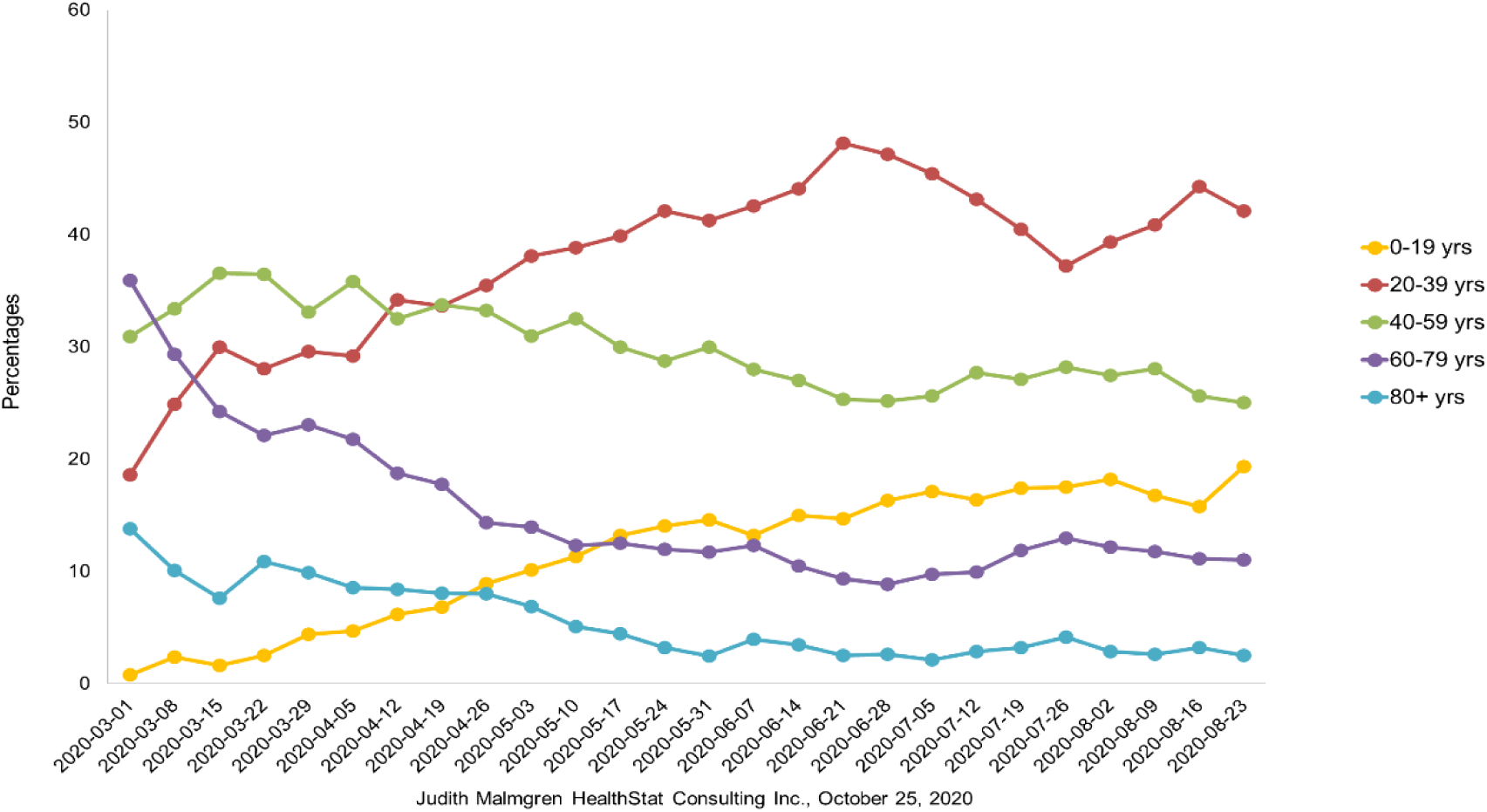
WA State DOH Confirmed Cases by age 3/1/2020-8/23/2020

## Discussion

Although morbidity and mortality from COVID-19 infection is highest among the infected population age 60 years and older and those with underlying conditions, the percentage of 0-19 and 20-39-year-old cases in Washington State increased over time with no decline in COVID-19 incidence among 0-19-year-olds and 20-39-year-olds compared to older age groups when stay-at-home orders were enacted at the beginning of the epidemic. Our results show that a persistently high percentage of current infections in Washington State exist among the age 0-19 and 20-39 population who may also be at highest risk of contracting and spreading the virus but not at high risk of hospitalization or mortality. During the first time period from March to April 2020, COVID-19 related hospitalization rates declined without an equivalent rate of decline among confirmed COVID-19 cases.

The shift from older to younger population COVID-19 infection illustrates the absence of a true decline in cases in the first time period and the extended need for health care capacity as the currently infected portion of the population is younger, less likely to report symptoms, at less risk of a severe life-threatening disease requiring hospitalization but fully capable of spreading disease to older, vulnerable adults. It appears persons 60 and older, those with pre-existing conditions and nursing home residents in particular were a harbinger of the serious nature of the epidemic but emphasis on infection in older adults only created a false dichotomy of risk at the beginning between young and old age. In Washington State 53% of the population is age 0-39 years, 25% 0-19 years and twenty eight percent aged 20-39 with Seattle/King County, the most populous county in the state.^8^

From August 2 to September 5, 2020, weekly COVID-19 cases among persons aged 18–22 years increased 55% nationally with greatest increases in the Northeast (144%) and Midwest (123%). Increases in cases were not solely attributable to increased testing.^9^ In Germany from March to April 2020, Goldstein et al report an observed relative increase of COVID-19 cases over time in 15-34 year olds compared to younger and older age groups.^10^ In South Korea, people age 20-39 numerically led the epidemic although the elderly suffered the majority of morbidity and mortality.^11^ In the southern United States regional outbreaks June 2020, positive COVID-19 cases among 20-39 year olds preceded spread to older age groups with percentage of cases among 0-39 year olds increasing over time (May-August 2020).^12^ Even though the largest percentage of cases are among younger age groups in the U.S., less than .3% of deaths are among 0-17 year olds and 1.9% of deaths are among 18-39 year olds.^13^ In Washington State the percentage of deaths by age are similar with zero deaths 0-19 years of age and 2% of total deaths 20-39 years of age.^14^ However, in a clinical profile of 3222 hospitalized young adults age 18-34 years in the U.S., this age group of patients did experience substantial rates of adverse outcomes although mortality was low compared to older age groups at 2.7%.^15^

As initial public warnings in Washington State and nationally targeted the population age 60 and older and those with underlying conditions, a misconception may have been at large that only persons age 60 and older were at risk for contracting COVID-19. Children were thought to be at low risk for COVID-19 and if infected to have a mild clinical course with few or no symptoms. However, a health alert was issued early in the epidemic by the CDC for Pediatric Multisystem Inflammatory System (MIS-C) linked to COVID-19 indicating serious morbidity risk exists although rare.^16, 17, 18, 19^ Documented outbreaks in WA State since going to Phase 2 reopening include childcare, K-12 schools, and college/university settings.^20^

Findings from a survey by Canning et al, found persons under age 50 had twice the predicted number of close contacts than older people.^21^ In a recent Center for Disease Control (CDC) study of persons age 18-23, social interactions, workplace and community transmission contributed to the sharp increase in cases among young adults after a ‘Safer at Home Emergency Order’ was invalidated in the state of Wisconsin.^22^ Misinformation and conflicting messaging influenced acceptance of and adherence to mitigation efforts. In another recent CDC survey study of engagement in mitigation behaviors, including handwashing, mask wearing and six-foot social distancing was lowest in those aged 18-29 years of age.^23^ The CDC urges colleges and universities to actively promote healthy environments.^24^ Although children and younger adults may not be at significant risk for serious COVID-19 related morbidity and mortality, evidence supports their ability to transmit disease and more likely occurrence of asymptomatic transmission endangering others they come in contact with especially older adults and those with comorbid conditions. In a CDC outbreak investigation, a single 13 year old index case was traced to 11 subsequent cases.^25^ A larger study of SARS-COV-2 infection in households in two states found when the index patient was less than age 12 secondary infection rate was 53% and age 13-17 years secondary infection rate was 38% with 75% of secondary infections occurring within a 5 day window of index case symptoms.^26^ From modelling studies it is estimated only 21% of infected children age 10-19 years exhibit symptoms.^27^ In a large cohort study in South Korea, 70% of 0-19, 63% of 20-29 and 60% of 30-39 year olds were asymptomatic.^28^ In a study by Lee et al of SARS-CoV-2 molecular viral shedding of symptomatic and asymptomatic patients, virus shedding was similar between the two groups.^29^ Limitations: While testing overall increased during the period evaluated, the percentage of younger people being tested decreased slightly compared to persons age 60 and older. Absent information to link family and household level data with parent’s occupation to identify the source of exposure in the 0-19-year-old age group, one could assume some infections are driven by contact with family members working outside of the home and social interaction as Washington State public schools have been closed since March 11, 2020. A study by the American Academy of Pediatrics found only 10 states reported testing by age further hampering the ability to evaluate level of COVID-19 disease incidence in the population by age.^30^ Testing by age was only available for Washington State in the first time period of our analysis. The broad age categories of 0-19 years and 20-39 years used by WA State DOH do not allow for more specific analysis regarding transmission and mitigation recommendations. Age of COVID-19 confirmed case positive is not reported by race in weekly online DOH data.

The Washington State stay-at-home policy is temporally associated with the first observed case decline after March 22, 2020.^31^ However the decline was only among age 40 and above with no decline in children or younger adults and cases rose again to a second peak in June indicating a persistent transmission rate among the population less than age 40 resistant to public health mitigation efforts. After mask wearing became required in all public spaces in Washington State (June 8, 2020) another decline in cases was observed without a decline in the percentage of cases among children and younger adults all while schools in Washington State remained closed.^32^

We document a shift of the majority of COVID-19 cases from older adults to children and younger adults over the eight-week time period after the state reached enough measurable cases to analyze on March 1, 2020. Further tracking of cases by age revealed a trend to majority COVID-19 infection among children and younger adults which continued upward to a sustained plateau regardless of fluctuations in statewide infection rates. Our findings indicate justifiable concern regarding the health and safety of children and teenagers that led to the decision to delay reopening schools. The disproportionate spread of disease among younger adults influenced plans for phased reopening of Washington State counties which are paused at Phase II for most counties until reduced case incidence and disease control testing, tracking and tracing metrics are met.^33^

With the shift in COVID-19 incidence to a majority of current cases in the combined 0-19/20-39-year-old age group statewide, counties with a high percentage population age 0-19 and 20-39 years are more heavily affected. Continued tracking of age trends are warranted and will help target which activities can be opened safely and what extra safeguards and protocols will be required with monitoring for adherence. Specific advisories tailored to children and adolescents age 0-19 years and 20-39-year-old adults to increase awareness of COVID-19 transmission and infection are advisable to reduce overall COVID-19 incidence levels and enhance movement towards levels that will allow a continuation of phased reopening of the state and counties. Adults age 20-39 specifically are more often employed in work sectors with high levels of public contact, more likely to be socially active and less likely to take social distancing and mask wearing precautions. Mitigation by low technical interventions will continue to be a priority for young adults and children even after a vaccine is available include mask wearing, social distancing, hand washing, adequate ventilation, and crowd avoidance.^34^ Modelling studies with real data simultaneously incorporating race and county with more precise age categories would be a tremendous asset to targeting specific public health mitigation strategies. Vaccination priority will be for medical workers and vulnerable persons by age or condition first with children and young adults lower priority creating a continued if not elevated need for specific mitigation while we await full population COVID-19 vaccination achievement.

## Supporting information

STROBE statement checklist

## Data Availability

Data downloaded October 1, 2020. All data is publicly available at
https://www.doh.wa.gov/Emergencies/COVID19/DataDashboard

## Acknowledgements

The authors wish to express their heartfelt thanks and gratitude for the invaluable assistance of Michelle Holshue, RN, MPH of the Washington State Department of Health and the CDC Epidemiologic Intelligence Service. The authors also wish to thank the Public Health staff in every county in Washington State and the Washington State Department of Health for their hard work collecting the data and their contribution to the Washington Disease Reporting System.

## Funding

No funding of any kind was received by any parties involved in this study.

All authors meet the conditions of authorship, 1) substantial contributions to conception and design, acquisition of data, or analysis and interpretation of data; 2) drafting the report or revising it critically for important intellectual content; and 3) final approval of the version to be published.

